# Preferences and patterns of response to public health advice during the COVID-19 pandemic

**DOI:** 10.1101/2021.02.15.21251765

**Authors:** Oded Nov, Graham Dove, Martina Balestra, Katharine Lawrence, Devin Mann, Batia Wiesenfeld

## Abstract

With recurring waves of the Covid-19 pandemic, a dilemma facing public health leadership is whether to provide public advice that is medically optimal (e.g., most protective against infection if followed), but unlikely to be adhered to, or advice that is less protective but is more likely to be followed. To provide insight about this dilemma, we examined and quantified public perceptions about the tradeoff between (a) the stand-alone value of health behavior advice, and (b) the advice’s adherence likelihood. In a series of studies about preference for public health leadership advice, we asked 1,061 participants to choose between (1) strict advice that is medically optimal if adhered to but which is less likely to be broadly followed, and (2) relaxed advice, which is less medically effective but more likely to gain adherence - given varying infection expectancies. Participants’ preference was consistent with risk aversion. Offering an informed choice alternative that shifts volition to advice recipients only strengthened risk aversion, but also demonstrated that informed choice was preferred as much or more than the risk-averse strict advice.

## Introduction

With recurring waves of the Covid-19 pandemic, preventing infection is a key public health strategy that frequently utilizes targeted advice about the behavior expected of the public. Public adherence to practices advised by health experts, however, has been uneven. Both high efficacy and high adherence are necessary for public health policies to generate desired outcomes, yet advice that most effectively protects people from infection (e.g., wearing a mask when outside one’s home) is often onerous, leading to low adherence. Public interventions ranging in strictness have been practiced since the early days of the pandemic (1-4). A dilemma facing public health leadership is whether to provide public advice that is medically optimal (e.g., most protective against infection if followed), but unlikely to be adhered to, or advice that is less protective but is more likely to be followed.

To provide insight about this dilemma, we examine and quantify public perceptions about the tradeoff between (a) the stand-alone value of health behavior advice (e.g. its efficacy in protecting against infection), and (b) the advice’s adherence likelihood (see Figures 3, 4). Following the scientific evidence supporting mask wearing (5, 6), in a series of studies we asked 1,061 participants to indicate their opinion about the appropriate public health advice strategies to be used by public health leadership, given different infection rates (i.e., the product of advice’s efficacy and adherence with it).

## Results

### Study 1

Using one-proportion z-tests with respect to the ‘indifference’ level (50%) within each condition we found that there was a significant preference for the strict advice strategy (N_strict_ = 51, 75% [64.7%, 85.3%]) over the relaxed advice (N_relaxed_=17, 25% [14.7%, 35.3%]) when the projected infection rates for ‘strict advice’ were lower than for ‘relaxed advice’ (z=4.761, p=0.000). However, when infection rates for strict advice were higher than for relaxed advice, there was no significant preference for either advice type (z=0.948,p=0.34); N_relaxed_=40, 56.3% [ 44.8%, 67.9%]; N_strict_=31, 43.7% [ 32.1%, 55.2%]) (see Figure 1).

**Figure 1.**
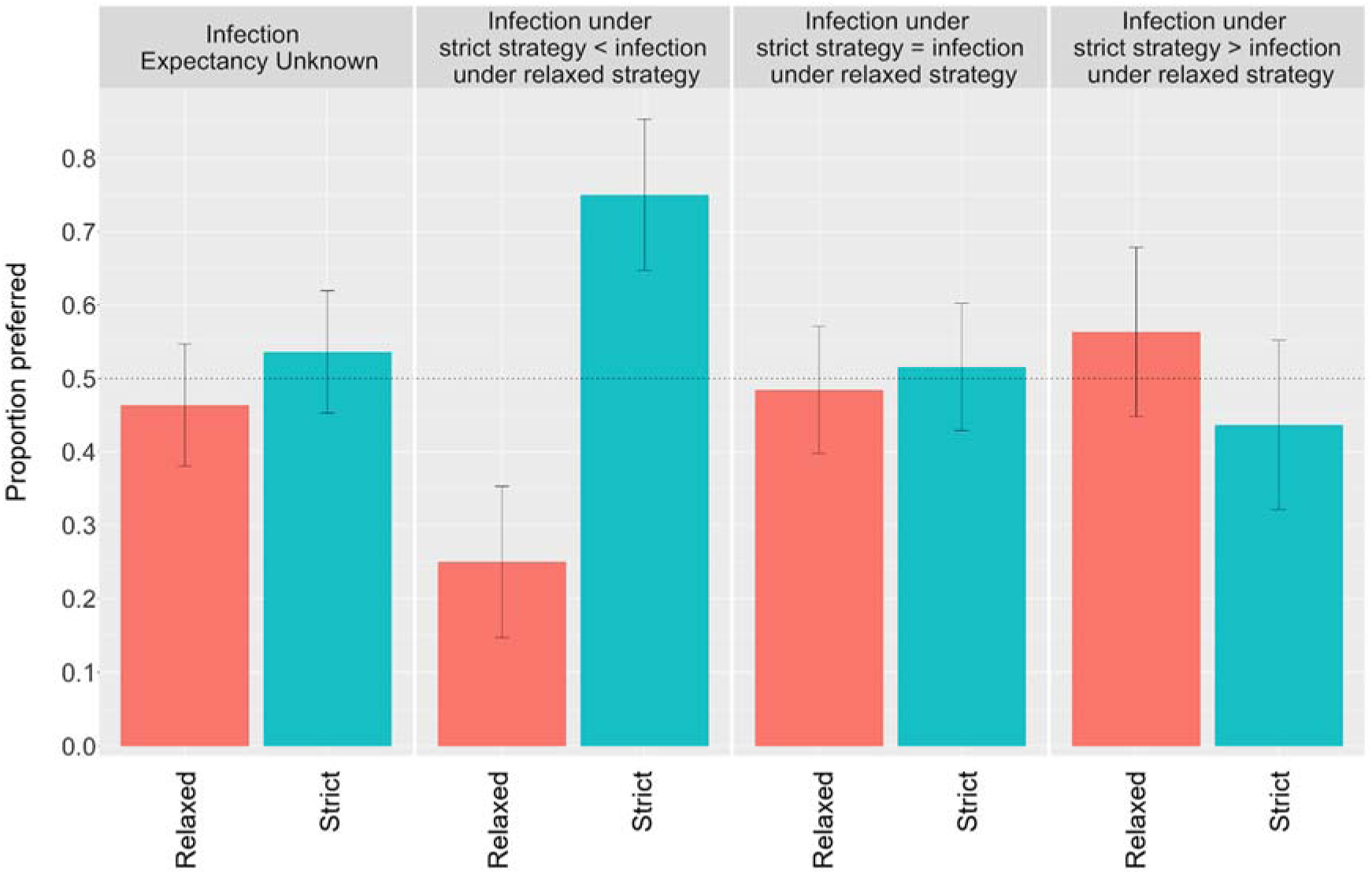
Study 1 results: Proportions of people who preferred each type of advice strategy under various infection expectancy conditions; dashed line: indifference rate (random choice between 2 options); error bars: 95% confidence intervals.

There was also no significant preference for advice type when infection expectancies were unknown (z=0.854, p=0.39; N_strict_=74, 53.6% [45.3%, 61.9%], (N_relaxed_=64, 46.4% [38.06%, 54.70%]) or equal (z=0.354, p=0.72; N_strict_=66, 51.6% [42.9%, 60.2%]; N_relaxed_=62, 48.4% [39.8%, 57.1%]).

### Study 2

A Chi-squared test of independence was performed to evaluate the relationship between the infection expectancy and participants’ preferred advice. The differences between infection expectancies on advice preference was significant (X-squared = 27.307, df = 6, p-value = 0.0001). Chi-squared goodness of fit tests were used to test if the distribution of preferences differed significantly from chance (i.e. probability of ⅓ of selecting any of the advice types) in each condition. Participants’ preferences were significantly different from chance in all four conditions (see Figure 2 for Chi-squares by condition).

**Figure 2.**
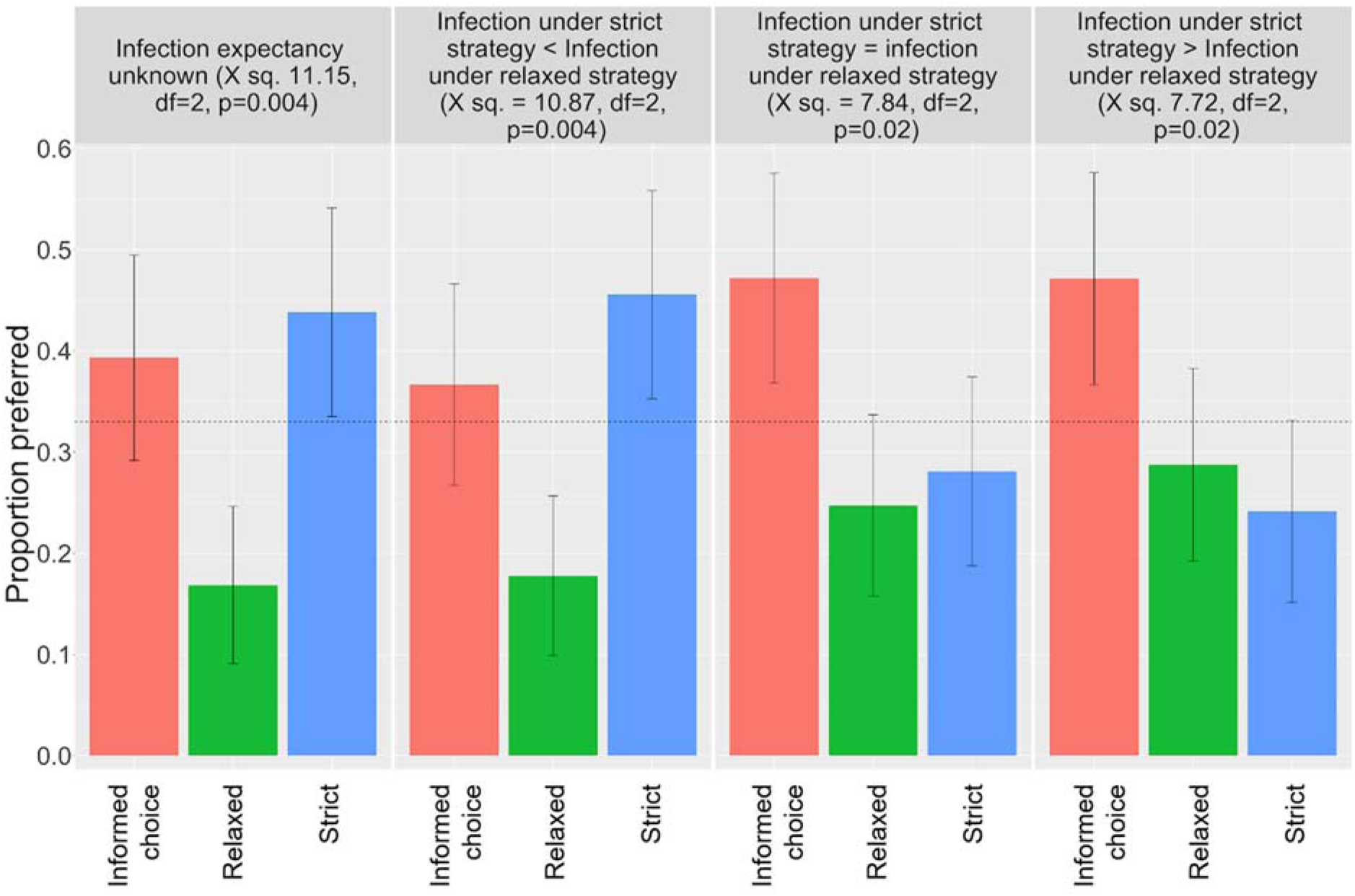
Study 2 results: Proportions of people who preferred each type of advice strategy under various infection expectancy conditions; dashed line: indifference rate (random choice between 3 options); error bars: 95% confidence intervals.

Across conditions, participants displayed a negative preference for the relaxed advice. When infection expectancies were unknown or higher for relaxed than strict advice, participants’ preference for the relaxed advice was significantly lower (N=15 (16.85% [9.08%, 24.63%]) and N=16 (17.78 [9.88, 25.68]), respectively) than for the strict advice (N=39 (43.82% [33.51%, 54.13%]) and N=41 (45.56% [35.27%, 55.84%]), respectively) or the informed choice conditions (N= 35 (39.32% [29.18%, 49.47%]) and N=33 (36.67% [26.71%, 46.62%]), respectively). When infection rates were equal for strict and relaxed advice, their preference for the relaxed advice (N=22 (24.72% [15.76%, 33.68%])) was significantly lower than the informed choice strategy (42 (47.19% [36.82%, 57.56%])). Even when infection rates indicated that the relaxed advice was superior, advice recipients merely became indifferent.

## Discussion

As public health leadership around the world considers different approaches to Covid-19 mitigation (7) in the face of differing perceptions and knowledge of the pandemic (8, 9) we offer insights into the perceived tradeoff between public health advice characteristics. Across Studies 1 and 2, where participants displayed a preference between alternative directive (relaxed vs. strict) advice, their preference was consistent with risk aversion. Offering an informed choice alternative that shifts volition to advice recipients only strengthened risk aversion, but also demonstrated that informed choice was preferred as much or more than the risk-averse strict advice.

Participants’ risk aversion suggests that they preferred policy makers to minimize social risk with stricter policies. A number of different values may drive this preference. Strict policies may be valued because of moral or ethical reasons, such as the notion that it is ethically desirable to prioritize prosocial (i.e., community-level well-being) over more self-interested (i.e., individual freedom) objectives when lives are at stake (10). Efficiency may also justify a preference for stricter policy advice; public risk, such as that associated with community transmission of a virus, is relatively inescapable and cannot be controlled by individual decision makers. People can therefore maximize their utility by reducing public risk that they cannot manage while accepting controllable private or personal risk, even perhaps in violation of strict directives.

The results of Study 2 suggest that informed choice, which combines information disclosure and recipient participation in decision-making, is preferred by advice recipients to policy maker control, particularly when information suggests that strict policies are not superior from an infection control perspective. Informed choice advice is both more informative and it offers greater freedom and volition to individuals, who have better knowledge of their own preferences. It could therefore be viewed as both more individually efficient and fairer. On the other hand, it is more ambiguous and uncertainty-provoking. In our studies, the differences in infection expectancies were not very large and the strict directive may not have been viewed as infringing on personal freedom much more than the relaxed directive. Preferences may shift more if these differences were larger.

While our work offers insights concerning public health guidelines in the face of a pandemic, it comes with a number of limitations: first, additional demographic data, such as level of education would have help understand preference patterns within the population. Second, participants were recruited from Amazon Mechanical Turk, which is limited with respect to population representativeness, and specifically, was recently found to skew higher towards males (11).

Taken together, the findings of both studies suggest that potential advice recipients prefer policy makers to reduce risk and enable informed choice, with a negative preference to a relaxed advice approach under any infection rate scenario.

## Methods

All experimental protocols were approved by NYU Institutional Review Board (#IRB-FY2020-4671). All methods were carried out in accordance with relevant guidelines and regulations. An informed consent was obtained from all subjects.

### Study 1

We recruited a total of 1,061 US-based Amazon Mechanical Turk (MTurk) participants who completed over 100 prior HITs with an approval rate greater than 95%. Participants could take part in this study - across all its variants - only once. Of these participants, 409 self-identified as female, 623 as male, 3 as non-binary, and the remainder did not respond. Their self-reported ages ranged between 18 and 78, with a mean age of 36.1 (median = 34, compared with a median age of 38 among the US population). Participants were paid a flat rate of either $2.50 for completing the study variants in which information about expected infection rates were provided (median time spent on these variants of the study was 3 minutes 24 seconds), or $2 for completing study variants in which information about expected infection rates were not provided (median time spent on these variants of the study was 5 minutes 14 seconds). This difference was due to the extended time required to read each of the explanatory paragraphs and infection rate examples.

Participants were given an attention check question, in which we asked them to identify the proportion of people that did not adhere to any advice who would be infected. This information was repeated in each of the explanatory text paragraphs, and so could be easily answered without reference to outside knowledge. Such attention checks are commonly used to address potential concerns over data validity in studies where participants are recruited through MTurk. We did not include the attention check question in study variants that did not include infection rate information. For these participants we relied solely on the more basic attention check of filtering for minimum time spent on the study (one minute) to exclude those participants who had simply rushed through the study without thought or attention. This inclusion criterion was also applied to participants who had passed the attention check question.

Participants were presented information on infection expectancies and asked to choose between two directive public health advice strategies:

A. Strict advice that is medically optimal for preventing infection if adhered to, but which is less likely to be broadly followed.
B. More relaxed advice, which is less medically effective at the individual level if adhered to, but more beneficial to reducing infection rates than no advice at all, and which will have higher rates of public adherence.

For the purpose of this study, ‘strict advice’ was described as ‘Advice that is medically best for preventing infection’, and participants were shown the example ‘Wear a mask at all times outside the home. Keep six feet away from others’. ‘Relaxed advice’ was described as ‘Useful advice that is more likely to be adopted by a larger proportion of people’, and the example ‘Wear a mask in enclosed public spaces where it is not possible to maintain a safe distance from others’. Participants were given additional explanatory text and an example of projected infection rates for a city of 10,000 inhabitants (see additional information in the “Text presented to participants” section). Participants’ preference – the outcome measure – were measured using a single item question in which a discrete choice was made by participants between options A and B.

To study the effect of different infection expectancies on participants’ perceptions of public health advice strategies, in Study 1 participants were either given no information about infection expectancies, or placed in one of three conditions describing infection expectancies associated with strict vs. relaxed advice (see Figure 3). They were then asked to indicate whether public health leadership should offer ‘strict advice’ or ‘relaxed advice’.

**Figure 3.**
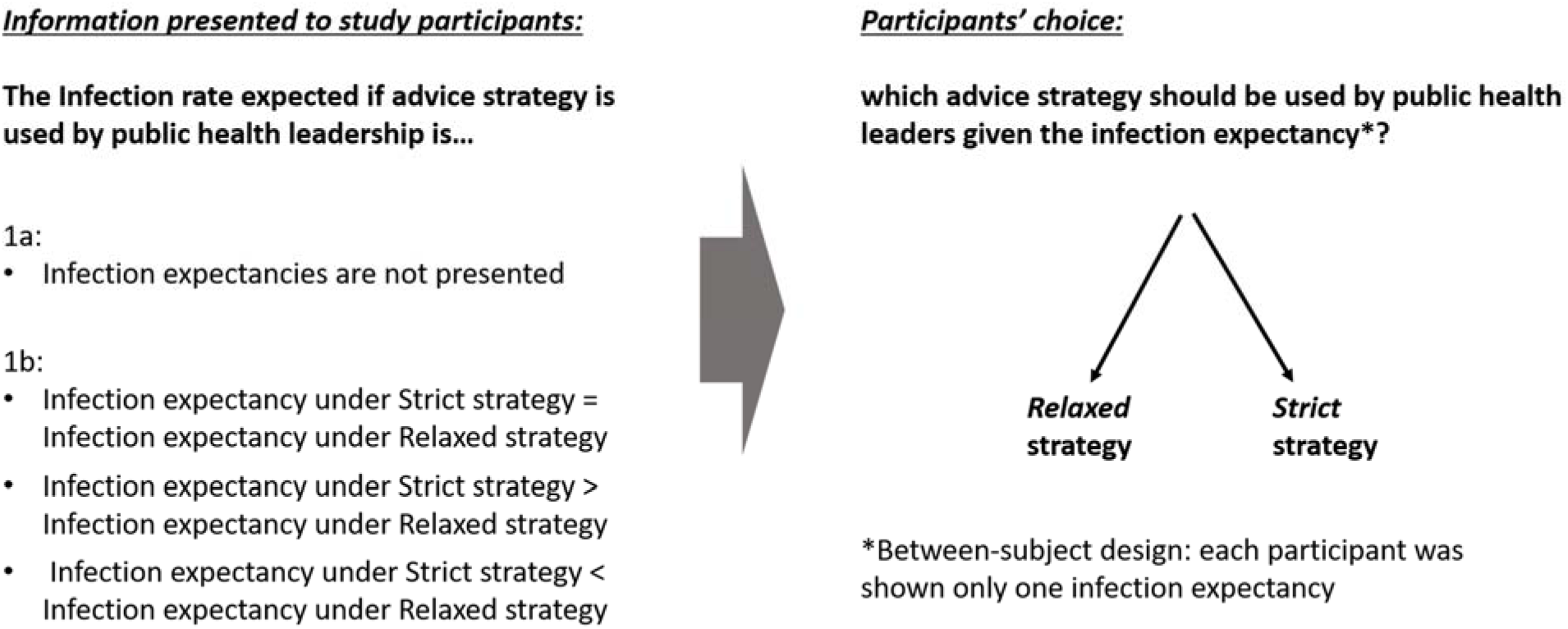
Experiment design, Study 1.

### Study 2

In study 2 (see Figure 4), we sought to evaluate the directive advice strategies (strict and relaxed) in relation to a strategy of informed decision-making (12) whereby the public is informed about the risks, benefits and acceptable alternatives, to empower them to make decisions taking their individual preferences into account. A third option was provided to participants to choose from, in addition to (A) and (B):

**Figure 4.**
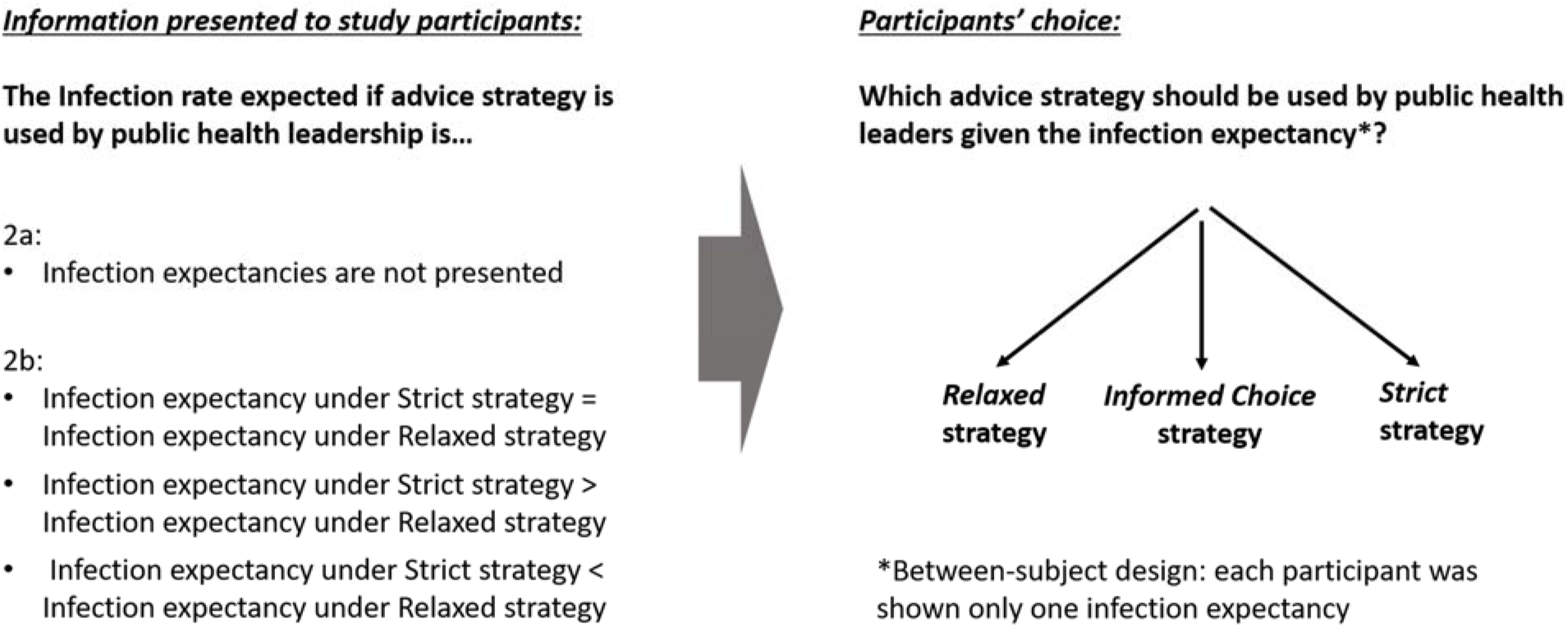
Experiment design, Study 2.

C. A strategy in which advice recipients (i.e., members of the public) are presented *both* advice types outlined in (A) and (B), along with their likely adherence and infection rates. (see additional information in the Supporting Information section).

With three options for participants to choose from, we varied the projected infection rates randomly so that for roughly one third of participants the total projected infection rates for presenting relaxed advice (1,420 out of the 10,000 city residents in the example text) and the informed choice advice (1,435) were lower than that for strict advice (1,525) (N=90), for another third of participants they were exactly equal (1,525) (N=89), and for the final third of participants the projected infection rates for relaxed advice (1,630) and informed choice advice (1,605) were higher than that for strict advice (1,525) (N=90). We again randomized the order in which the strict and relaxed advice options appeared on the screen. As in Study 1, participants’ preference – the outcome measure – were measured using a single item question in which a choice was made by participants between options A, B, C.

### Text presented to participants

Text presented to participants in each study variant, reflecting the different conditions - adherence rates expected infection rates variations:

#### Strict advice

Advice that is medically best for preventing infection.

“Wear a mask at all times outside the home. Keep 6 feet away from others.”

- Among people who comply with this advice there is an average infection rate of 1% within 1 month, and on average 25% of the public are likely to comply with this advice. The infection rate among the remainder who do not comply with this advice is 20% within 1 month. Example: In a city with a population of 10,000 people, it is likely that 2,500 will comply with this advice and on average 25 of these will be infected. The remaining 7,500 people are likely not to comply with this advice, and on average 1,500 of these will be infected. In this scenario, it is likely that a total of 1,525 people out of the city’s population of 10,000 will be infected within 1 month.

#### Relaxed advice

Useful advice that is more likely to be adopted by a larger proportion of people.

“Wear a mask in enclosed public spaces where it is not possible to maintain a safe distance from others.”

**Three expected infection variations:**

- Among people who comply with this advice there is an average infection rate of 10% within 1 month, and on average 47.5% of the public are likely to comply with this advice. The infection rate among the remainder who do not comply with this advice is 20% within 1 month. Example: In a city with a population of 10,000 people, it is likely that 4,750 will comply with this advice and on average 475 of these will be infected. In contrast, the remaining 5,250 are likely not to comply with this advice, and on average 1,050 of these will be infected. In this scenario, a total of 1,525 people out of the city’s population of 10,000 will be infected within 1 month.
- Among people who comply with this advice there is an average infection rate of 10% within 1 month, and on average 37% of the public are likely to comply with this advice. The infection rate among the remainder who do not comply with this advice is 20% within 1 month. Example: In a city with a population of 10,000 people, it is likely that 3,700 will comply with this advice, and on average 370 of these will be infected. The remaining 6,300 are likely not to comply with this advice, and on average 1,260 of these will be infected. In this scenario, a total of 1,630 people out of the city’s population of 10,000 will be infected within 1 month.
- Among people who comply with this advice there is an average infection rate of 10% within 1 month, and on average 58% of the public are likely to comply with this advice. The infection rate among the remainder who do not comply with this advice is 20% within 1 month. Example: In a city with a population of 10,000 people, it is likely that 5,800 will comply with this advice, and on average 580 of these will be infected. The remaining 4,200 are likely not to comply with this advice, and on average 840 of these will be infected. In this scenario, a total of 1,420 people out of the city’s population of 10,000 will be infected within 1 month.

#### Informed choice

Offer the public both types of advice.

“The best medical advice is to wear a mask at all times outside the home and keep 6 feet away from others. However, wearing a mask in enclosed public spaces where it is not possible to maintain a safe distance from others, also offers some protection from infection.”

Three expected infection variations:

- If both sets of advice are offered, we would expect reduced adherence with each in comparison to when offered alone. Among people who comply with the best medical advice there is an average infection rate of 1% within 1 month, and on average 15% of the public are likely to comply with this advice. Among people who comply with the advice more likely to be adopted there is an average infection rate of 10% within 1 month, and on average 37% of the public are likely to comply with this advice. The infection rate among the remainder who do not comply with either advice is 20% within 1 month. Example: In a city with a population of 10,000 people, it is likely that 1,500 will comply with the medically-best advice, and on average 15 of these will be infected. A further 1,900 are likely to comply with the advice more likely to be adopted, and on average 190 of these will be infected. The remaining 6,600 are likely not to comply with either advice, and on average 1,320 of these will be infected. In this scenario, a total of 1,525 will be infected within 1 month.
- If both sets of advice are shown we would expect reduced adherence with each in comparison to when offered alone. Among people who comply with the best medical advice there is an average infection rate of 1% within 1 month, and on average 15% of the public are likely to comply with this advice. Among people who comply with the advice more likely to be adopted there is an average infection rate of 10% within 1 month, and on average 11% of the public are likely to comply with this advice. The infection rate among the remainder who do not comply with either advice is 20% within 1 month. Example: In a city with a population of 10,000 people, it is likely that 1,500 will comply with the medically-best advice, and on average 15 of these will be infected. A further 1,100 are likely to comply with the advice more likely to be adopted, and on average 110 of these will be infected. The remaining 7,400 are likely not to comply with either advice, and on average 1,480 of these will be infected. In this scenario, a total of 1,605 will be infected within 1 month.
- If both sets of advice are shown we would expect reduced adherence with each in comparison to when offered alone. Among people who comply with the best medical advice there is an average infection rate of 1% within 1 month, and on average 15% of the public are likely to comply with this advice. Among people who comply with the advice more likely to be adopted there is an average infection rate of 10% within 1 month, and on average 28% of the public are likely to comply with this advice. The infection rate among the remainder who do not comply with either advice is 20% within 1 month. Example: In a city with a population of 10,000 people, it is likely that 1,500 will comply with the medically-best advice, and on average 15 of these will be infected. A further 2,800 are likely to comply with the advice more likely to be adopted, and on average 280 of these will be infected. The remaining 5,700 are likely not to comply with either advice, and on average 1,140 of these will be infected. In this scenario, a total of 1,435 will be infected within 1 month.

## Data Availability

All data is available upon request

## Acknowledgments

This work was supported by the National Science Foundation award #1928614.

